# Coinfection Ecology and Pathogen Emergence in a *Borrelia*-Endemic Landscape: Five Years of *Borrelia burgdorferi, Anaplasma phagocytophilum*, and *Babesia microti* Surveillance in Maryland

**DOI:** 10.64898/2025.12.03.25341558

**Authors:** Greg Joyner, Olifan Abil, Maria J. Sanches, Amy Schwartz, Julia Poje, Kathryn Arnold, Christine Petersen, Maria Gomes Solecki

**Affiliations:** Department of Microbiology, Immunology and Biochemistry, University of Tennessee Health Science Center, Memphis, Tennessee, USA; Immuno Technologies Inc, Memphis, Tennessee, USA; University of Iowa, Iowa City, IA, USA

## Abstract

The emergence of tick-borne pathogens depends on ecological opportunity and barriers to persistence within vectors and hosts. *Borrelia burgdorferi* is firmly entrenched in the mid-Atlantic, whereas *Babesia microti* and *Anaplasma phagocytophilum* remain patchily distributed. Five years of integrated surveillance (2020–2024) at three Maryland sites allowed us to track *B. microti* and *A. phagocytophilum* establishment by screening questing *Ixodes scapularis* nymphs, *Peromyscus*-fed nymphs, and *Peromyscus leucopus* by qPCR, then contextualizing results with paired county-level human case data. *B. burgdorferi* was consistently detected in all sites and sample types, with prevalence stable at approximately 5–20% in questing nymphs and exceeding 30% in hosts, confirming long-term enzootic maintenance. By contrast, *B. microti* and *A. phagocytophilum* were initially sporadic but increased in prevalence, particularly in rodents and *Peromyscus*-fed ticks. Over time *A. phagocytophilum* prevalence significantly increased to above 20% in some *Peromyscus*-fed nymphal collections despite much lower prevalence in questing ticks, highlighting the early-warning value of bloodmeal-associated surveillance. Coinfections were rare, though enrichment of *B. burgdorferi* + *A. phagocytophilum* in *Peromyscus*-fed ticks suggests possible facilitation during early establishment. These results indicate that *B. microti* and *A. phagocytophilum* are actively emerging in Maryland, following their entrenchment in the Northeast and Upper Midwest. Combining surveillance from questing nymphal ticks, *Peromyscus*-fed nymphal ticks, and *P. leucopus* reservoir hosts provides a framework for detecting enzootic cycles before they appear in questing populations or human case counts, offering critical early-warning capacity for public health preparedness.

**IMPORTANCE:** Understanding why some tick-borne pathogens become ecosystem entrenched while others remain sporadic is central to predicting human disease emergence. By combining surveillance of questing nymphal *Ixodes* ticks, *Peromyscus*-fed nymphal ticks, and *Peromyscus leucopus* reservoir hosts across five years in Maryland, we show that *Babesia microti* and *Anaplasma phagocytophilum* remain in the early stages of ecological entrenchment whereas *Borrelia burgdorferi* is deeply established. This integrated approach demonstrates how pathogen biology within the tick shapes field prevalence and highlights *Peromyscus*-fed ticks as a powerful xenodiagnostic early-warning tool for detecting emerging pathogens before they are reflected in questing populations or human case data.

## INTRODUCTION

Lyme disease, caused by *Borrelia burgdorferi,* is deeply ecologically established in the northeastern and mid-Atlantic United States. In contrast, *Babesia microti* (babesiosis) *and Anaplasma phagocytophilum* (anaplasmosis) have more recently emerged as enzootic cycles in these regions. The broader category of tick-borne diseases continues to expand its geographic and public health impact (1, 2), with *Anaplasma* and *Babesia* posing new challenges for surveillance and control due to their complex ecology, varied transmission dynamics, and potential for asymptomatic infection in humans (3–8).

The enzootic cycle of *B. burgdorferi* is characterized by high reservoir competence in *Peromyscus leucopus*, efficient acquisition by *Ixodes scapularis*, and consistent vector-to-host transmission (9–12). In contrast*, B. microti* and *A. phagocytophilum* display distinct patterns of prevalence, persistence, and transmission efficiency, particularly in emerging regions (13, 14). *Babesia microti* is a protozoan parasite with a two-host life cycle involving obligate vertebrate and arthropod stages. Infection in rodents is typically asymptomatic and can persist for months, providing a sustained source of transmission to feeding larvae (5, 15). Ticks must acquire *B. microti* as larvae to be infectious as nymphs, as transstadial passage is required and transovarial transmission does not occur (11, 12).

*A. phagocytophilum* is an obligate intracellular bacterium that replicates within neutrophils in vertebrate hosts and within midgut or salivary tissues in the tick vector. Enzootic maintenance depends primarily on larval acquisition, with nymphs serving as the main transmission stage; thus, most nymphal infections reflect prior larval acquisition (6, 10, 16, 17). However, nymphal acquisition is biologically possible, though rarely demonstrated experimentally, and would generate infected adults, which may contribute to human risk even if this pathway plays little role in long-term persistence. Field detections of *A. phagocytophilum* in adult *I. scapularis* have been observed (18, 19). Transovarial transmission has not been demonstrated (17, 20, 21).

Coinfections involving *B. burgdorferi* and *B. microti* are increasingly recognized in clinical settings and may contribute to more severe or prolonged illness in immunocompromised patients (22–26). In contrast, coinfections with *A. phagocytophilum* are less consistently associated with exacerbated clinical outcomes, and current evidence does not suggest a clear augmentative effect between *A. phagocytophilum* and other tick-borne pathogens (27). In fact, there is evidence that *A. phagocytophilum* may reduce the inflammatory response induced by *B. burgdorferi*. (Scorza et al, Inf. Immune). Nonetheless, such coinfections remain diagnostically important, especially in transitional regions where ecological signals may precede clinical awareness.

At the ecological level, coinfection of *Ixodes scapularis* with multiple pathogens has been documented across endemic and transitional zones, though mechanisms remain incompletely understood. Shared hosts and synchronized seasonal dynamics may contribute to coinfection, but there is also speculation that the presence of one pathogen might facilitate acquisition or persistence of another. For example, *B. microti* prevalence in ticks often lags but tracks that of *B. burgdorferi*, potentially reflecting the use of established host–vector systems during regional expansion (28–32).

Rather than seeking to establish first detection, this study aims to describe the trajectory of *B. microti* and *A. phagocytophilum* emergence in Maryland, where entrenchment appears to be ongoing. In this context, emergence refers not merely to pathogen DNA detections but to ecological establishment - sustained transmission, multi-year persistence, and increasing prevalence in both ticks and competent reservoir hosts. Understanding these differences is essential not only for anticipating emergence but also for designing targeted surveillance strategies. Our approach builds on previous calls for long-term, integrated surveillance across both hosts and vectors to better understand interannual dynamics (2, 4, 9, 33).

Historical studies in endemic regions such as southern New York and Nantucket Island demonstrated stable cycles of *B. microti* in rodents and nymphs decades ago, yet similar entrenchment only recently extended further south, often after decades of lag relative to *B. burgdorferi* (18). Surveillance of blood donors and hospitalized patients has revealed increasing cases of babesiosis in previously low-incidence states, including a >1,400% rise in case numbers in Maine and Vermont between 2011 and 2019 (34). A recently published study evaluating *Ixodes* across the eastern seaboard indicated emergence of *Babesia microti* in Maryland and Delaware between 2013 and 2023, also finding coinfected ticks (35). Maryland lies at a critical latitudinal and ecological transition between historically endemic zones (e.g., New York, Pennsylvania) and regions where *B. microti* and *A. phagocytophilum* have only recently become detectable. Its climatic convergence with the Northeast, along with forest fragmentation and dense *Peromyscus* populations, make it an ideal sentinel zone for tracking ecological penetrance (31, 36).

We present a five-year dataset (2020–2024) from three ecologically distinct sites in Maryland, each sampled consistently using drag and live-trapping protocols, and analyzed via qPCR for *B. burgdorferi*, *B. microti*, and *A. phagocytophilum*. Infection data from questing nymphs, host-attached nymphs, and *Peromyscus leucopus* hosts are synthesized alongside county-level human case reports to assess the degree of establishment. While coinfections were infrequent in this dataset, they remain epidemiologically relevant due to their potential to exacerbate disease severity and complicate clinical diagnosis, particularly in newly affected regions.

## METHODS

### Field sites and sampling

Fieldwork was conducted from May to July in each of five consecutive years (2020–2024) at three sites in Maryland, USA: Montgomery County (GFH; ∼39.28°N, −77.10°W), Harford County (EHH; ∼39.58°N, −76.52°W), and Baltimore County (HHB; ∼39.49°N, −76.83°W), as previously described (37). All sites were privately owned properties used for the breeding and housing of hunting hounds, situated within a rural–suburban landscape matrix of farmland, mixed woodland, and maintained grass fields. Each site covered approximately 40,000 m² (4 ha) with a mixture of mature forest, secondary growth, and ecotone habitats with trail systems and fencing throughout. The site in Baltimore county was enrolled in the study in 2022-2024.

Sampling alternated weekly from May to August between small mammal trapping and tick dragging. At each site, linear transects (>5 per site) were established for both trapping and dragging to ensure spatial coverage. For small mammals, Sherman live traps (20–40 per site per night, spaced at approximately 15 m intervals) were baited with chow and deployed overnight. Traps were checked each morning, non-target species were released immediately, and captured *Peromyscus leucopus* were briefly restrained by scruff for tick removal. Animals were released at the capture site without sedation or euthanasia.

Questing *Ixodes scapularis* nymphs were collected by dragging a 1 m² white flannel cloth across vegetation along woodland and edge transects. Drags were conducted during daylight hours and paused during periods of high heat or low humidity. The cloth was examined at approximately 15 m intervals, and ticks were removed with forceps, placed in labeled tubes, and stored at −20 °C. Sampling continued until a minimum threshold of nymphs was obtained per site per year, ensuring prevalence estimates could be made but not directly proportional to density.

### Tick identification and staging

All ticks were identified morphologically as *Ixodes scapularis* nymphs. Species identity was confirmed molecularly by PCR amplification of a 152 bp fragment of the actin gene using primers IxAct-F (5′-GCCCTGGACTCCGAGCAG-3′) and IxAct-R (5′-CCGTCGGGAAGCTCGTAGG-3′). Only confirmed nymphs were included in analyses; no larvae or adults were used.

### DNA extraction

Ticks were surface sterilized sequentially in 10% bleach, 70% ethanol, and sterile PBS, then homogenized individually in 0.5 mL bead-beating tubes prefilled with baked zirconia beads (Glen Mills, Cat. 7361-002000) using a Mini-Beadbeater-24 for 1.5 min. DNA was extracted with a modified Qiagen DNeasy Blood & Tissue Kit (Qiagen, Cat. 1017647) including Proteinase K and carrier RNA. Lysates were clarified by centrifugation (6,000 × *g*, 10 min), mixed with AL buffer and ethanol, and passed through spin columns with standard wash steps (AW1, AW2, ethanol). DNA was eluted twice in 50 µL AE buffer and stored at −20 °C. Negative extraction controls were included once per batch.

### Pathogen detection

*Borrelia burgdorferi* was detected using a TaqMan qPCR targeting the flaB gene (forward primer: 5′-AAGCAATCTAGGTCTCAAGC-3′; reverse primer: 5′-GCTTCAGCCTGGCCATAAATAG-3′; probe: 5′-FAM-AGATGTGGTAGACCCGAAGCCGAG-TAMRA-3′). Reactions were run in 20 µL volumes on a QuantStudio 7 Real-Time PCR System (Applied Biosystems) with cycling conditions of 95 °C for 10 min, followed by 45 cycles of 95 °C for 15 s and 60 °C for 1 min. Samples were considered positive if amplification was equivalent to ≥50 flaB copies per sample, based on standard curves generated from cultured *B. burgdorferi* DNA.

*Babesia microti* and *Anaplasma phagocytophilum* were detected using Taqman Multiplex Real-Time PCR Assay (Thermo Scientific reagents) targeting 18S rRNA and 16S rRNA genes, respectively. *Babesia microti* 18S rRNA primers (forward5′-GCATGGAATAATGAAGTAGGACTTTGGT-3′ and reverse 5′-CCCCAACTGCTCCTATTAACCATT-3′) and a probe (5’-[FAM]-CTCTGGCTCAATAACC –[MGB]-3); *Anaplasma phagocytophilum* 16S primers (forward 5′CGGAATTCCTAGTGTAGAGGTGAAA-3′ and reverse 5′-GTCAGTACCGGACCAGATAGC-3′) and a probe (5’-[VIC]-CCACTGGTGTTCCTCC-[MGB]-3’). PCR amplification was performed in a 20 µL reaction mixture containing 2 µL of total DNA extract, 900 nM of each primer, and 250 nM of probe. Cycling conditions were 95 °C for 20 s, followed by 40 cycles of 95 °C for 1 s and 60 °C for 20 s. The TaqMan multiplex real-time PCR assay was optimized by first performing both single plex and multiplex reactions under identical conditions, using the same reagents and run simultaneously on the same plate. If the cycle threshold (Ct) values obtained from the single plex and multiplex reactions were comparable, subsequent analyses were performed using the multiplex PCR. All samples were tested in duplicate. A Ct ≤ 35 was considered positive. Melt-curve analysis was not performed. Synthetic gBlocks (Integrated DNA Technologies) corresponding to the complete target regions of the flaB, 18S rRNA, and 16S rRNA genes, together with field-confirmed positive samples, were included as assay controls. For assay validation, 20 positive samples per pathogen were reamplified with long-range primers (>400 bp) and sequenced on an Oxford Nanopore platform, confirming species identity.

### Data analysis

Samples were scored positive if amplification occurred within defined thresholds. Coinfection was defined as simultaneous detection of ≥2 pathogens in the same tick. Prevalence and 95% Wilson confidence intervals were calculated per site and year. Differences in prevalence by site, year, and tick source (drag vs. *Peromyscus*-fed) were tested using Fisher’s exact test or chi-squared tests. Multiple-testing correction was applied with Bonferroni and FDR adjustments using the statsmodels package (v0.14.2). All analyses were performed in Python v3.12.7.

### Ethics statement

All animal handling followed institutional and federal guidelines. *Peromyscus leucopus* were captured with live traps, examined without sedation, and released after tick removal and submandibular bleeding. Tick removal and blood sampling were performed by trained personnel. IACUC protocol number 25-0643.

### Human case data

County-level anaplasmosis and babesiosis case counts for 2014–2023 were obtained from the Maryland Department of Health (MDH) (38). Data were aggregated by county and year. Population denominators were taken from U.S. Census Bureau annual estimates to calculate incidence rates. Case definitions followed MDH confirmed/probable classifications. All data used were publicly reportable and contained no personally identifiable information.

## RESULTS

### Questing nymphal infection prevalence

We evaluated the prevalence of *Borrelia burgdorferi, Babesia microti*, *and Anaplasma phagocytophilum* in questing nymphal *Ixodes scapularis* collected by drag sampling across three Maryland field sites from 2020 to 2024 (Figure 1A). *B. burgdorferi* was consistently detected at all sites, with prevalence ranging from 5–20%. This stable background underscores its long-term entrenchment in Maryland, likely following introduction from more established Northeast foci during the 1980s, as *I. scapularis* expanded its range southward (39–41). At Harford County, prevalence fluctuated but remained detectable each year, from 6.5% in 2023 to 21.4% in 2021. Montgomery County followed a similar pattern, with low prevalence in 2020 (5.3%) rising to 17.1% by 2024. In Baltimore County, *B. burgdorferi* was nearly absent in 2022 (2.0%) but increased to 17.4% in 2023 and 13.5% in 2024, despite fewer ticks overall.

**Figure 1.**
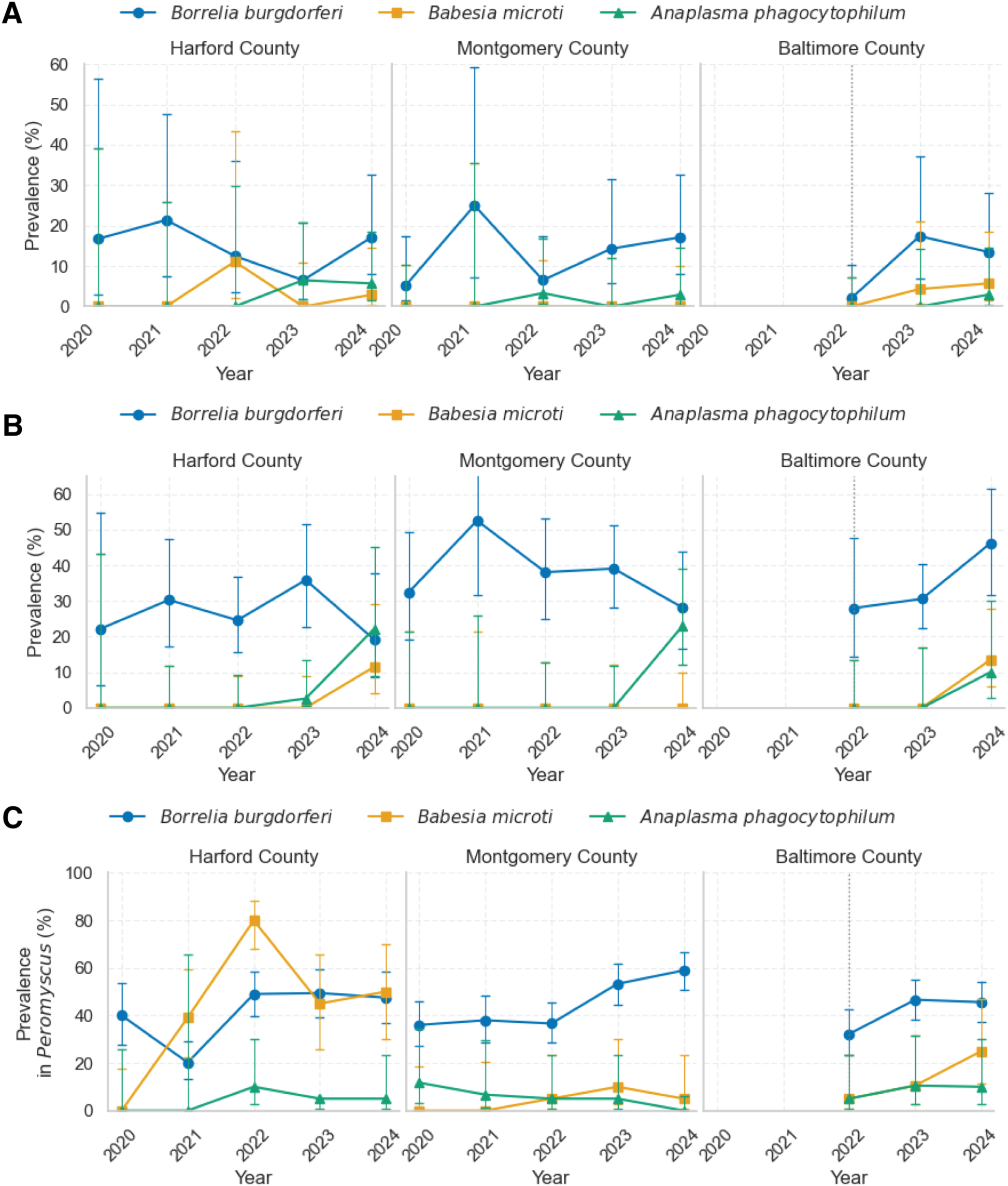
Prevalence of *Borrelia burgdorferi*, *Babesia microti*, and *Anaplasma phagocytophilum* across three field sites in Maryland (2020–2024): (A) Flat questing nymphal *Ixodes scapularis*. (B) Engorged nymphs collected from trapped *Peromyscus leucopus*. (C) *Peromyscus leucopus* hosts. Each site is shown in a separate panel. Prevalence (%) was calculated as the proportion of individuals testing positive for each pathogen by qPCR. For (C), *B. burgdorferi* prevalence was determined by ear tissue qPCR, while *B. microti* and *A. phagocytophilum* prevalence were determined by blood qPCR. Error bars represent 95% confidence intervals (Wilson’s method).

By contrast, *B. microti* and *A. phagocytophilum* were only sporadically detected and always at low prevalence. In Harford County, *B. microti* first appeared in 2022 (11.1%, 1/9 positives) and recurred at 2.9% in 2024, while *A. phagocytophilum* was detected in both 2023 (6.5%) and 2024 (5.7%). Montgomery County showed an emerging *A. phagocytophilum* signal in 2022 (3.3%) and 2024 (2.9%), but no *B. microti* across the study period. In Baltimore County, *B. microti* appeared in 2023 (4.3%) and 2024 (5.7%), while *A. phagocytophilum* was detected only once (2024, 2.9%).

Logistic regression adjusting for site suggested a modest, non-significant increase in *B. microti* prevalence over time (odds ratio [OR] per year = 1.96, 95% CI 0.67–5.76, p = 0.22). *A. phagocytophilum* showed a stronger indication of rising prevalence (OR per year = 2.15, 95% CI 0.89–5.22, p = 0.089). Pooled chi-square tests for trend did not detect significant monotonic increases (*B. microti* p = 0.60; *A. phagocytophilum* p = 0.41). Together, these results highlight the patchy, stochastic nature of early emergence, with occasional site- and year-specific detections rather than smooth linear increases.

### Mouse-fed nymph infection prevalence

Infection prevalence in *Peromyscus*-fed nymphs (Figure 1B) was consistently higher than in questing ticks, particularly for *B. burgdorferi*. Across sites and years, *B. burgdorferi* prevalence typically ranged from 20–50%, about 2–3× higher than in drag-collected nymphs (compare with Figure 1A). This increase reflects repeated opportunities for acquisition across both the larval and nymphal bloodmeal.

At Harford county, *B. burgdorferi* prevalence fluctuated between 19.2% (2024) and 35.9% (2023), broadly tracking host infection levels observed in Peromyscus. At Montgomery county, prevalence reached 52.6% in 2021 and remained consistently high in subsequent years (28–39%). At Baltimore County*, B. burgdorferi* prevalence increased steadily from 28.0% in 2022 to 46.2% in 2024, suggesting a strong and persistent local enzootic cycle.

Signals for *Babesia microti* and *Anaplasma phagocytophilum* were more sporadic but revealed higher prevalence in *Peromyscu*s-fed ticks than in questing ticks, especially in later years. *B. microti* appeared in Harford County (2024: 11.5%) and Baltimore (2024: 13.5%) but was absent at the site in Montgomery County. By contrast, *A. phagocytophilum* showed a clear emergence in both Harford (2024: 22.2%) and Montgomery (2024: 22.9%), while Baltimore County yielded lower but nonzero detections (2024: 10.0%).

Notably, A. phagocytophilum prevalence in Peromyscus-fed nymphs was nearly an order of magnitude higher than in questing ticks, exceeding 20% in some site–years (Figure 1B). At the Harford County site (EHH), logistic regression revealed a statistically significant annual increase (OR = 8.81, 95% CI: 1.23–63.03, p = 0.0302). A combined statewide model across all sites showed an even stronger effect (OR = 16.18, 95% CI: 2.37–110.67, p = 0.0045), supporting active expansion of *A. phagocytophilum* within rodent-associated tick populations. These patterns indicate that *A. phagocytophilum* is circulating robustly in rodent hosts and detected readily in feeding nymphs before it becomes apparent in the questing population, consistent with an early, patchy stage of establishment.

### Combined trend analysis across ticks

When data from both questing and Peromyscus-fed nymphs were analyzed together, strong evidence emerged for increasing prevalence of *B. microti* and *A. phagocytophilum* over time. Logistic regression adjusting for site and collection method indicated significant year-on-year increases: *B. microti* prevalence rose approximately 4.7-fold per year (OR = 4.7, 95% CI 1.6–13.7, p = 0.004), while *A. phagocytophilum* rose approximately 5.4-fold per year (OR = 5.4, 95% CI 2.3–12.5, p < 0.001).

Pooled chi-square trend tests confirmed these increases (*B. microti* p = 0.0009; *A. phagocytophilum* p = 6.8 × 10⁻⁶). Site effects were consistent with the individual analyses: Harford and Montgomery counties generally showed higher *A. phagocytophilum* prevalence relative to Baltimore County. Collection method (questing vs. *Peromyscus-*fed) was not significant for *B. microti* but approached significance for *A. phagocytophilum,* consistent with stronger inference from *Peromyscus*-fed ticks.

### Peromyscus infection prevalence

Infections in *Peromyscus leucopus* provided the strongest evidence of enzootic circulation of all three pathogens, with prevalence (Figure 1C) consistently exceeding that observed in ticks.

*B. burgdorferi* remained entrenched, with prevalence generally ranging from 30–60%. In Harford County, prevalence was stable at approximately 40–50% across 2020–2024. Montgomery County increased from 36.0% in 2020 to 58.9% in 2024. Baltimore County, sampled only from 2022 onwards, showed stable prevalence between 32.2% and 46.6%.

*B. microti* prevalence in hosts was more variable, with sharp peaks in Harford County (80.0% in 2022, 50.0% in 2024). This transient surge in 2022 may represent a founder event or stochastic amplification. Montgomery County showed only sporadic positives (≤10%), while Baltimore County demonstrated gradual increases (5.0% in 2022 to 25.0% in 2024).

*A. phagocytophilum* emerged sporadically but was generally detected earlier in hosts than in ticks. At Harford County, prevalence rose from absent in 2020–2021 to 10.0% in 2022, peaking at 22.2% in 2024. Montgomery County showed intermittent low-level infection (5.0% in 2022–2023) before dropping to zero in 2024. Baltimore County showed consistent detections (5–10%) from 2022–2024. Importantly, *A. phagocytophilum* prevalence in hosts was detected in several site–years where questing nymph prevalence was zero, highlighting rodent surveillance as a leading indicator of pathogen establishment.

### Coinfection analysis

Coinfections among *Ixodes scapularis* nymphs were rare (Table 1). Most ticks carried a single pathogen, and coinfection combinations occurred only sporadically.

**Table 1.** Observed and Expected Coinfections in *Ixodes scapularis* Ticks by Collection Method: Comparison of observed coinfection frequencies of *Borrelia burgdorferi* (Bb), *Anaplasma phagocytophilum* (Ap), and *Babesia microti* (Bm) in drag ticks vs ticks from *Peromyscus* across three Maryland field sites (2020–2024).

| Coinfection Type | Method | n | Observed Count | % of Ticks | Expected Count | Fisher's Exact p |
| --- | --- | --- | --- | --- | --- | --- |
| **Borrelia + Anaplasma** | Drag | 333 | 2 | 0.60% | 0.82 | 0.192 |
| *Peromyscus*-fed | 332 | 9 | 2.71% | 5.02 | **0.046** |  |
| **Borrelia + Babesia** | Drag | 333 | 0 | 0.00% | 0.59 | 1.000 |
| *Peromyscus*-fed | 332 | 5 | 1.51% | 2.67 | 0.123 |  |
| **Anaplasma + Babesia** | Drag | 333 | 0 | 0.00% | 0.11 | 1.000 |
| *Peromyscus*-fed | 332 | 1 | 0.30% | 0.36 | 0.312 |  |
| **Triple (Bb + Ap + Bm)** | Drag | 333 | 0 | 0.00% | 0.01 | N/A |
| *Peromyscus*-fed | 332 | 1 | 0.30% | 0.12 | N/A |  |

The only statistically significant enrichment was *B. burgdorferi* + *A. phagocytophilum* coinfections in *Peromyscus*-fed nymphs, detected in 9 ticks (2.6%) compared to an expected 4.8 under independence (p = 0.023). This enrichment may reflect ecological facilitation via shared hosts or overlapping seasonal transmission windows.

Other coinfection types were either absent or occurred at chance levels. *B. burgdorferi* + *B. microti* coinfections were detected twice in *Peromyscus*-fed ticks and not at all in questing ticks. *A. phagocytophilum* + *B. microti* coinfections were not observed. A single triple coinfection (*B. burgdorferi* + *A. phagocytophilum* + *B. microti*) was detected in a single *Peromyscus*-fed nymph but was too rare for statistical analysis.

### Human case trends of emerging tick-borne diseases

Confirmed human cases of anaplasmosis and babesiosis in Maryland remain rare compared with northeastern hotspots, but both have increased gradually over the past decade (Figure 2A–B; Supplementary Table 2; Maryland Department of Health, 2025 (38)). From 2014–2018, statewide reports of both diseases were sporadic, typically ≤3 cases per year, and many counties reported none. Beginning in 2019, however, a clearer upward trajectory emerged for both pathogens, with repeated detections in central and western Maryland.

**Figure 2.**
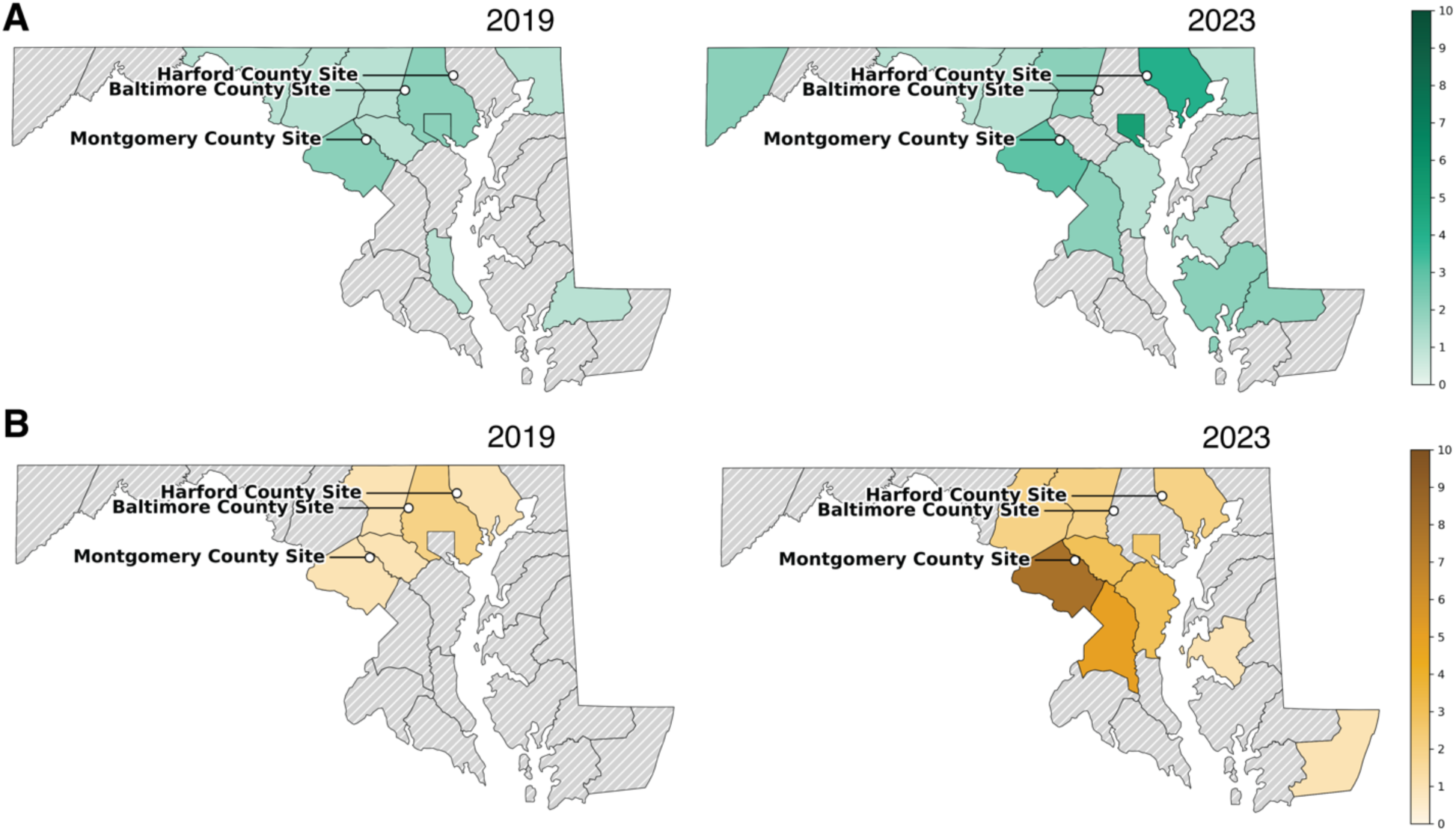
Human anaplasmosis and babesiosis incidence in Maryland: Shown are annual confirmed human cases by county for two representative years from the study period: 2019 (left) and 2023 (right). (A) Human anaspasmosis cases. (B) Babesiosis cases. Case data were obtained from Maryland Department of Health surveillance records.

For anaplasmosis, statewide case counts rose steadily to a peak of 29 cases in 2022–2023. Montgomery County displayed the most consistent signal, with cases increasing from 1–2 annually in the mid-2010s to 6 cases in both 2021 and 2022. Harford County reported occasional cases in the late 2010s and rose to 4 in 2023, while Baltimore City showed a sharp increase from isolated detections in 2017 to 5 cases in 2019 and 4 in 2023. By contrast, Baltimore County reported low-level cases earlier in the decade but none after 2019.

Babesiosis showed a similar but slightly delayed pattern. From 2014–2018, annual statewide totals were ≤3, but by 2023 cases had increased to 27. Montgomery County again showed the strongest signal, rising from isolated detections in 2018–2019 to 8 confirmed cases in 2023, the highest annual total for any Maryland county. Harford County reported its first case in 2018 with additional detections in 2019 and 2023, while Baltimore County recorded one or two cases in multiple years between 2016 and 2022.

Together, these patterns indicate that both *A. phagocytophilum* and *Babesia microti* are in the process of ecological establishment in Maryland, with Montgomery County appearing to be at the forefront of increasing human incidence. Although absolute numbers remain low compared with the northeastern United States, the concordance between rising case counts and our enzootic surveillance signals suggests that local transmission cycles are strengthening.

## DISCUSSION

### Emergence in a Borrelia-endemic landscape

Predicting pathogen emergence in transitional zones requires surveillance methods that detect early signals before pathogens become entrenched in questing tick populations. In Maryland, *Borrelia burgdorferi* is long established, but our five-year surveillance reveals increasing yet incomplete establishment of *Babesia microti* and *Anaplasma phagocytophilum*. The strongest signals appear in *Peromyscus* hosts and *Peromyscus*-fed nymphs, indicating active but still developing enzootic cycles.

### Biological constraints and contrasting establishment

*B. burgdorferi* remains entrenched due to its efficient persistence in the tick midgut and regulated migration to salivary glands during feeding (42–44). In contrast, *A. phagocytophilum* must colonize midgut and hemocytes and manipulate tick immune processes (45–51), creating multiple bottlenecks that may explain low prevalence in questing ticks even when host infection is frequent.

*B. microti* must undergo sexual development in the tick after acquisition and lacks transovarial transmission (11, 12, 29). This dependency on reacquisition each generation, combined with reduced metabolic flexibility (52, 53), yields inefficient transmission and contributes to patchier establishment. These life-cycle differences help explain why *B. microti* and *A. phagocytophilum* initially emerge more strongly in hosts and feeding ticks than in questing ticks.

### *Peromyscus*-fed nymphs as early-warning indicators

In multiple site-years, *A. phagocytophilum* prevalence in *Peromyscus*-fed nymphs exceeded 20%, nearly order-of-magnitude higher than in questing ticks. Some detections likely reflect DNA in the bloodmeal rather than stable tick tissue infection (54). Mammalian permissiveness, including immune dampening in dogs and mice (6, 27), may support high host infection without efficient vector persistence. Regardless of mechanism, these findings demonstrate that *Peromyscus*-fed ticks capture early pathogen circulation that would be underestimated using drag sampling alone.

Rising Maryland human cases from 2019–2023, especially in Montgomery County, reinforce that *Peromyscus*-fed tick surveillance can serve as a leading indicator of establishment.

### Coinfection dynamics during early establishment

Coinfections were rare overall, consistent with incomplete emergence of *B. microti* and *A. phagocytophilum*. The only statistically significant association was enrichment of *B. burgdorferi* + *A. phagocytophilum* coinfections in *Peromyscus*-fed nymphs. Conversely, *B. burgdorferi* + *B. microti* coinfections, widely documented in ecological (14, 49, 55) and clinical studies (26, 56, 57), were scarce. One hypothesis is that early establishment of *A. phagocytophilum* overlaps more directly with entrenched *B. burgdorferi* cycles in *Peromyscus*, whereas *B. microti* arrival is more recent and stochastic. The absence of *A. phagocytophilum* + *B. microti* combinations may reflect suppressive dynamics observed in experimental models (15, 29, 58).

### Public health implications

Although absolute Maryland case numbers remain low, both anaplasmosis and babesiosis show clear upward trends. *B. microti* is the leading transfusion-transmitted parasite in the United States (59–61), and *A. phagocytophilum* can cause severe disease in high-risk populations (7, 27, 62). The concordance of rodent, tick-fed, and human case trends suggests strengthening local transmission and highlights the need for proactive surveillance.

### Regional context and ecological drivers

Emergence patterns in Maryland mirror historical trajectories in the Northeast and Upper Midwest, where *B. microti* and *A. phagocytophilum* lagged decades behind *B. burgdorferi* establishment (12, 18, 21, 63, 64). Environmental factors including land-use change, deer abundance, mast cycles, and climate warming shape tick and pathogen dynamics (65–68). As a transitional ecotone adjoining long-entrenched northeastern foci, Maryland appears poised for continued pathogen spread southward and westward.

### Limitations and future directions

This study did not characterize pathogen genotypes nor confirm tissue-established tick infection. Priority next steps include integrating strain-level analyses, confirmation of salivary gland colonization particularly for *A. phagocytophilum* ecotypes (36), and expansion across the broader mid-Atlantic to assess geographic generalizability.

## Conclusion

*B. burgdorferi* remains deeply established in Maryland, whereas *B. microti* and *A. phagocytophilum* are clearly emerging. *Peromyscus*-fed ticks provide an early-warning signal that precedes widespread establishment in questing ticks and should be incorporated into routine surveillance. Integrated monitoring across hosts, fed ticks, and questing ticks will be essential for anticipating tick-borne disease emergence in transitional landscapes.

## Data Availability

All data produced in the present study are available upon reasonable request to the authors

## ACKNOWLEDGMENTS

We thank the field teams and property owners who supported tick and small-mammal collections in Harford, Baltimore, and Montgomery Counties. In particular, we acknowledge Paige Witucki and Eli Davis for their field assistance, as well as the broader field team members who contributed to collections and sample processing across the five-year study. We also thank Filipe Azevedo and Aseel Alamri for their contributions to laboratory processing and data support.

## SUPPLEMENTARY MATERIAL

**Supplementary Table 1.**
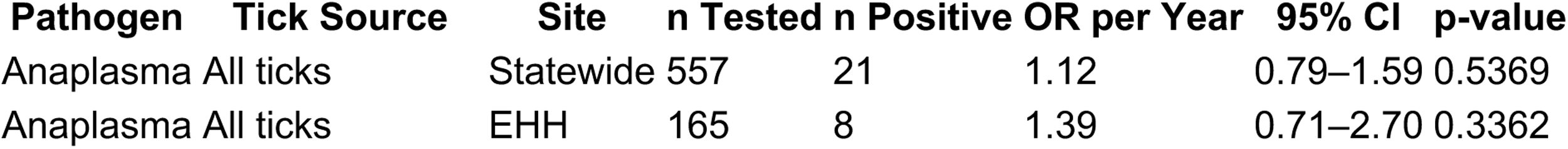
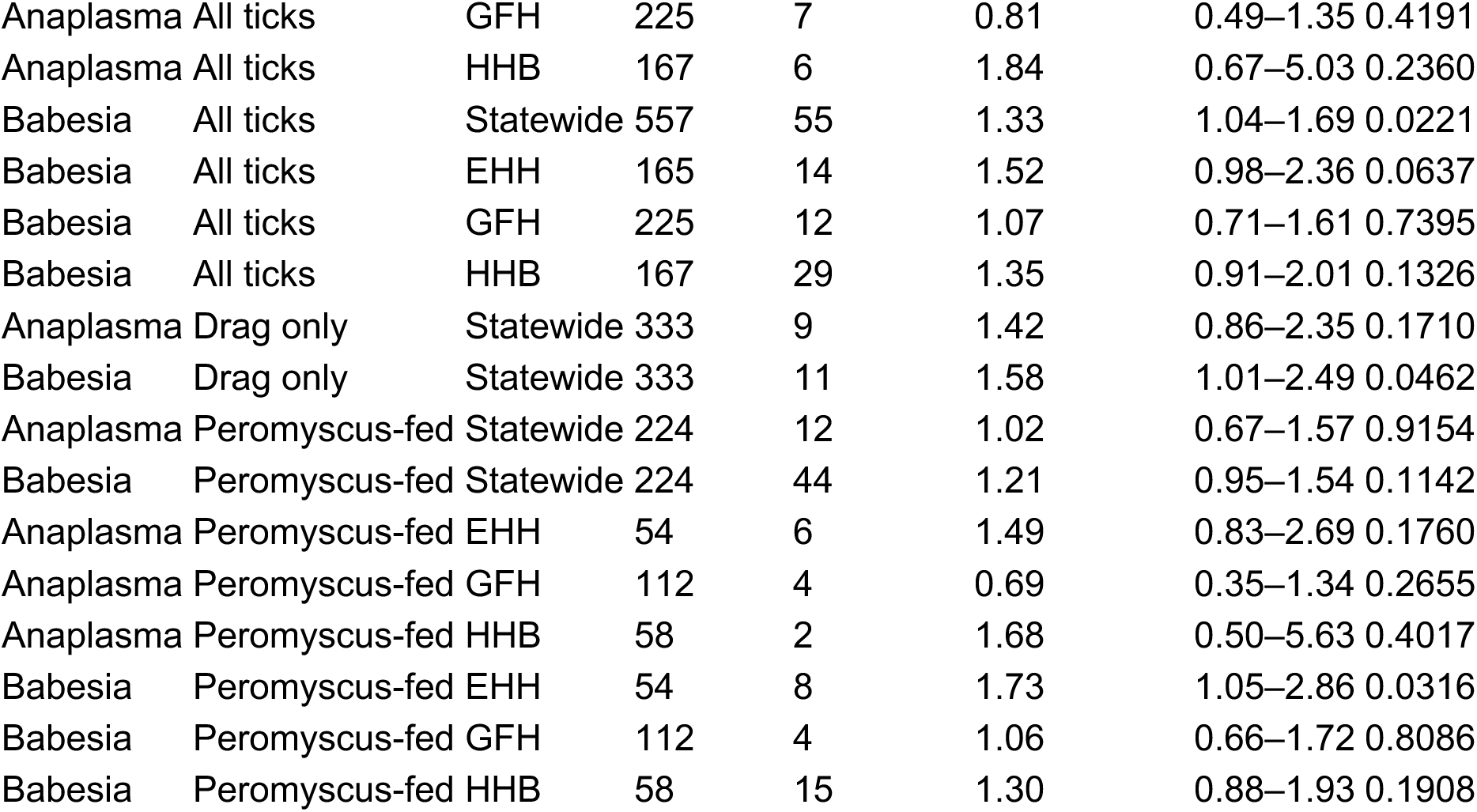
Logistic Regression of Infection Prevalence by Year: Logistic regression results testing for monotonic changes in infection prevalence over time (2020–2024), stratified by pathogen, tick type, and site. OR = Odds Ratio; CI = Confidence Interval.

| Pathogen | Tick Source | Site | n Tested | n Positive | OR per Year | 95% CI | p-value |
| --- | --- | --- | --- | --- | --- | --- | --- |
| Anaplasma | All ticks | Statewide | 557 | 21 | 1.12 | 0.79–1.59 | 0.5369 |
| Anaplasma | All ticks | EHH | 165 | 8 | 1.39 | 0.71–2.70 | 0.3362 |
| Anaplasma | All ticks | GFH | 225 | 7 | 0.81 | 0.49–1.35 | 0.4191 |
| Anaplasma | All ticks | HHB | 167 | 6 | 1.84 | 0.67–5.03 | 0.2360 |
| Babesia | All ticks | Statewide | 557 | 55 | 1.33 | 1.04–1.69 | 0.0221 |
| Babesia | All ticks | EHH | 165 | 14 | 1.52 | 0.98–2.36 | 0.0637 |
| Babesia | All ticks | GFH | 225 | 12 | 1.07 | 0.71–1.61 | 0.7395 |
| Babesia | All ticks | HHB | 167 | 29 | 1.35 | 0.91–2.01 | 0.1326 |
| Anaplasma | Drag only | Statewide | 333 | 9 | 1.42 | 0.86–2.35 | 0.1710 |
| Babesia | Drag only | Statewide | 333 | 11 | 1.58 | 1.01–2.49 | 0.0462 |
| Anaplasma | Peromyscus-fed | Statewide | 224 | 12 | 1.02 | 0.67–1.57 | 0.9154 |
| Babesia | Peromyscus-fed | Statewide | 224 | 44 | 1.21 | 0.95–1.54 | 0.1142 |
| Anaplasma | Peromyscus-fed | EHH | 54 | 6 | 1.49 | 0.83–2.69 | 0.1760 |
| Anaplasma | Peromyscus-fed | GFH | 112 | 4 | 0.69 | 0.35–1.34 | 0.2655 |
| Anaplasma | Peromyscus-fed | HHB | 58 | 2 | 1.68 | 0.50–5.63 | 0.4017 |
| Babesia | Peromyscus-fed | EHH | 54 | 8 | 1.73 | 1.05–2.86 | 0.0316 |
| Babesia | Peromyscus-fed | GFH | 112 | 4 | 1.06 | 0.66–1.72 | 0.8086 |
| Babesia | Peromyscus-fed | HHB | 58 | 15 | 1.30 | 0.88–1.93 | 0.1908 |

**Supplementary Table 2.** Confirmed Human Cases of Anaplasmosis and Babesiosis in Maryland by County, 2019–2023: Case counts were obtained from Maryland Department of Health public surveillance data.

| County | Anaplasmosis<br>2019 | 2020 | 2021 | 2022 | 2023 | Babesiosis<br>2019 | 2020 | 2021 | 2022 | 2023 |
| --- | --- | --- | --- | --- | --- | --- | --- | --- | --- | --- |
| Allegany | 0 | 0 | 0 | 0 | 0 | 0 | 0 | 0 | 0 | 0 |
| Anne Arundel | 0 | 1 | 3 | 1 | 1 | 0 | 0 | 0 | 0 | 3 |
| Baltimore City | 5 | 0 | 0 | 1 | 4 | 0 | 1 | 1 | 0 | 2 |
| Baltimore<br>County | 2 | 0 | 0 | 0 | 0 | 2 | 2 | 2 | 2 | 0 |
| Calvert | 1 | 0 | 0 | 2 | 0 | 0 | 0 | 0 | 0 | 0 |
| Caroline | 0 | 0 | 0 | 0 | 0 | 0 | 0 | 0 | 0 | 0 |
| Carroll | 1 | 0 | 2 | 1 | 2 | 1 | 0 | 2 | 1 | 2 |
| Cecil | 1 | 1 | 0 | 0 | 1 | 0 | 0 | 1 | 0 | 0 |
| Charles | 0 | 0 | 0 | 0 | 0 | 0 | 0 | 0 | 0 | 0 |
| Dorchester | 0 | 0 | 0 | 0 | 2 | 0 | 0 | 0 | 0 | 0 |
| Frederick | 1 | 0 | 1 | 0 | 1 | 0 | 0 | 2 | 0 | 2 |
| Garrett | 0 | 0 | 0 | 1 | 2 | 0 | 0 | 0 | 0 | 0 |
| Harford | 0 | 0 | 0 | 1 | 4 | 1 | 0 | 0 | 0 | 2 |
| Howard | 1 | 0 | 2 | 2 | 0 | 1 | 1 | 1 | 0 | 3 |
| Kent | 0 | 0 | 0 | 0 | 0 | 0 | 0 | 0 | 0 | 0 |
| Montgomery | 2 | 4 | 6 | 6 | 3 | 1 | 3 | 3 | 2 | 8 |
| Prince<br>George's | 0 | 0 | 0 | 1 | 2 | 0 | 1 | 0 | 0 | 5 |
| Queen<br>Anne's | 0 | 0 | 0 | 0 | 0 | 0 | 0 | 0 | 0 | 0 |
| St. Mary's | 0 | 0 | 0 | 0 | 0 | 0 | 0 | 0 | 0 | 0 |
| Somerset | 0 | 0 | 0 | 1 | 0 | 0 | 0 | 0 | 0 | 0 |
| Talbot | 0 | 0 | 0 | 0 | 1 | 0 | 0 | 0 | 0 | 1 |
| Washington | 1 | 0 | 0 | 0 | 1 | 0 | 0 | 0 | 0 | 0 |
| Wicomico | 1 | 0 | 1 | 0 | 2 | 0 | 0 | 0 | 1 | 0 |
| Worcester | 0 | 0 | 2 | 0 | 0 | 0 | 1 | 1 | 1 | 1 |

